# A simple direct RT-LAMP SARS-CoV-2 saliva diagnostic

**DOI:** 10.1101/2020.11.19.20234948

**Authors:** Michael J. Flynn, Olga Snitser, James Flynn, Samantha Green, Idan Yelin, Moran Szwarcwort-Cohen, Roy Kishony, Michael B. Elowitz

**Author notes:** Contact Information: Michael J. Flynn, Michael B. Elowitz.

## Abstract

Widespread, frequent testing is essential for curbing the ongoing COVID-19 pandemic. Because its simplicity makes it ideal for widely distributed, high throughput testing, RT-LAMP provides an attractive alternative to RT-qPCR. However, most RT-LAMP protocols require the purification of RNA, a complex and low-throughput bottleneck that has often been subject to reagent supply shortages. Here, we report an optimized RT-LAMP-based SARS-CoV-2 diagnostic protocol for saliva and swab samples. In the protocol we replace RNA purification with a simple sample preparation step using a widely available chelating agent, as well as optimize key protocol parameters. When tested on clinical swab and saliva samples, this assay achieves a limit of detection of 10^5^ viral genomes per ml, with sensitivity close to 90% and specificity close to 100%, and takes 45 minutes from sample collection to result, making it well suited for a COVID-19 surveillance program.

## Introduction

### Widespread testing is seen as the solution for preventing the spread of SARS-CoV-2, but must include the testing of asymptomatic people to be effective

While much progress has been made in addressing the COVID-19 pandemic, treatments remain limited and vaccines have not finished deployment. In the meantime, societies’ best option for containment is to follow “test and trace” approaches where those recently exposed to SARS-CoV-2 quarantine themselves so they do not spread the virus further. However, in most countries including the US, tests are typically administered only after symptoms develop. A growing scientific consensus holds that effective epidemiological approaches for containing the virus must also include routine testing of asymptomatic people^1^.

### Testing for SARS-CoV-2 after the onset of symptoms misses a crucial window of pre-symptomatic infectivity

Viral loads decrease over time in most patients hospitalized with SARS-CoV-2, indicating a peak in viral load sometime at or before symptom onset^2^. This is consistent with data from a nursing home outbreak showing a subpopulation of presymptomatic patients with very high viral loads (C_t_ ∼13.7-18) that was not seen in the symptomatic patients^3^. Similarly, statistical analysis on cases of transmission comparing time between symptom onsets with the incubation period of the virus inferred a peak infectivity 0-1 days before symptom onset^4^. If viral load and infectivity peak before symptom onset, then it is likely that a patient will have spread the virus before they are able to quarantine themselves. In this vein, modeling indicates that testing will only be effective if conducted on asymptomatic individuals across the population^1^. However, operating at this scale places significant constraints on the design of a test.

### The most common and effective testing method, RT-qPCR, suffers from high costs, a requirement for infrastructure and operational complexity

Polymerase chain reaction (PCR) is a foundational method in molecular biology whereby short DNA “primers” are annealed to a DNA template of interest, extended by DNA polymerase, creating a copy of the template, and then dissociated at high temperature to begin the cycle again. Quantitative polymerase chain reaction (qPCR) couples this DNA synthesis to fluorescence using a DNA binding dye or other method, which allows a quantitative readout. By comparing the number of cycles (doublings) required to reach a measurable fluorescence, the C_t_ value, the amount of template DNA in the sample can be roughly quantified. Reverse Transcriptase qPCR, RT-qPCR, adds a reverse transcription reaction to the beginning in order to target RNA. RT-qPCR is an essential laboratory method and highly sensitive, able to detect down to 10^3^ SARS-CoV-2 genome copies per ml in clinical swab samples. However, the assay also has several drawbacks we hope to address in this work. The first aspect to improve is cost: the price of a COVID-19 qPCR test is limiting in nearly all settings and ranges from $20 to greater than $100 per test. A second, related aspect is equipment to run the qPCR test, which can cost many thousands of dollars. A third aspect is test complexity: an RT-qPCR test requires molecular biology expertise to conduct and interpret, which restricts its execution to labs equipped with this expertise (CLIA labs in the USA). The resulting logistical challenge of collecting, shipping, and processing samples from different locations at these limited number of labs increases test turnaround time and reduces throughput. An ideal test would be cheap and simple enough to conduct at the point of care in low resource settings.

### Other limitations of the general testing regime include RNA extraction and sample collection

RT-qPCR and other methods require RNA extraction from a sample before use in the assay. RNA extraction is costly, low throughput, subject to reagent shortages, and requires molecular biology expertise to conduct, which makes it desirable to skip. Additionally, nasopharyngeal swabs have often been in short supply and are painful and dangerous to collect. By contrast, saliva collection, which is non-invasive, safer to collect, and not dependent on swabs or other supplies, yields comparable or higher viral loads compared to nasopharyngeal swabs^5^. Thus, an ideal assay would skip RNA extraction and could be run on saliva.

### A surveillance assay need not be as sensitive as qPCR

After infection is established, RNA viruses enter an exponential phase during which viral load rises from undetectable to orders of magnitude higher than the LOD of qPCR in as little as 1-2 days. Based on this fact, mathematical modeling suggests that diagnostic tests that have 1000-fold higher LOD than qPCR can still be effective, if individuals are tested often enough to catch the spike^1^. Therefore, sensitivity can be traded off to satisfy the other constraints.

### Colorimetric RT-LAMP can address many of the key issues currently limiting testing

Colorimetric Reverse Transcriptase Loop-mediated Isothermal Amplification (RT-LAMP) is a method for detecting a specific RNA species using an isothermal polymerase reaction. In this method, primers are designed such that a target RNA triggers a chain reaction of DNA hairpin synthesis^6,7^, which is read out by a pH sensitive dye that turns yellow as protons are released by DNA polymerization^8^. Since it does not require a PCR machine, RT-LAMP is ideal for field testing of viral infections and has been used for Ebola diagnosis and surveillance in Guinea^9,10^, for tracking Zika virus in Brazilian mosquito populations^11^, and for COVID-19 diagnosis during the initial outbreak in Wuhan and Shenyang, China^12,13^. Recent work has solidified RT-LAMP as a simple, inexpensive, and sufficiently sensitive alternative to RT-qPCR for SARS-CoV-2 detection^14–26^.

### Here, adding a chelating agent and optimizing reaction volume, we demonstrate significant improvements to RT-LAMP

First we show that the non-hazardous chelating agent Chelex-100 can be used to prepare saliva samples for RT-LAMP, similar to how it has been used to prepare complex forensic samples for PCR^27–29^. By sequestering divalent metal cations, Chelex-100 protects RNA from degradation at high temperature. Chelex-100 also has a basic pH which reduces sensitivity to variations in patient saliva pH without inhibiting LAMP. Second, we found that larger reaction volumes can provide greater sensitivity. With these optimizations, we achieved a limit of detection of 10^5^ RNA copies per ml and got results consistent with RT-qPCR on clinical swab samples with C_t_ up to 32, yielding a sensitivity near 90% and specificity near 100% on clinical swab and saliva samples.

## Results

### Chelex-100 can partially protect RNA from high temperature degradation

We first set out to eliminate the need for RNA purification from saliva samples. To create synthetic samples for testing, the SARS-CoV-2 N gene (IDT #10006625) was in-vitro transcribed, quantified with digital PCR, serially diluted in 10-fold steps, and spiked into human saliva. The samples could then be treated with various sample preparation techniques and added to a 10 µl colorimetric RT-LAMP reaction for readout, for which we used a previously published primer set targeting the N gene, N2^22^ (see Methods). We explored multiple approaches to inactivate potential inhibitors in the samples, including proteinase K to degrade protein-based inhibitors as well as a 95C heat inactivation step.

With heat inactivation, we confirmed a recently reported result that assay sensitivity was significantly higher when RNA is added after saliva has been heat treated than before, implying a significant amount of temperature-induced degradation^17^(Figure 1, second and third panels). We hypothesized that RNA degradation at high temperatures is caused by enhancement of RNA hydrolysis by divalent metal cations, which mediate nuclease activity at biological temperatures^30^. These cations attack phosphodiester bonds, cleaving the RNA backbone^30^. However, chelating agents can sequester these cations. In particular, the chelating agent Chelex-100 has been used to prepare RNA and DNA for PCR from complex forensic samples such as blood and saliva^28,29^. In keeping with standard Chelex-100 protocols, synthetic samples were mixed 2:3 with 10% wt/vol Chelex-100 solution, incubated at 95C for 10 minutes, and then assayed with RT-LAMP. Using this sample prep, the degradation due to heat inactivation was reduced 100-fold (Figure 1, fourth panel).

**Figure 1:**
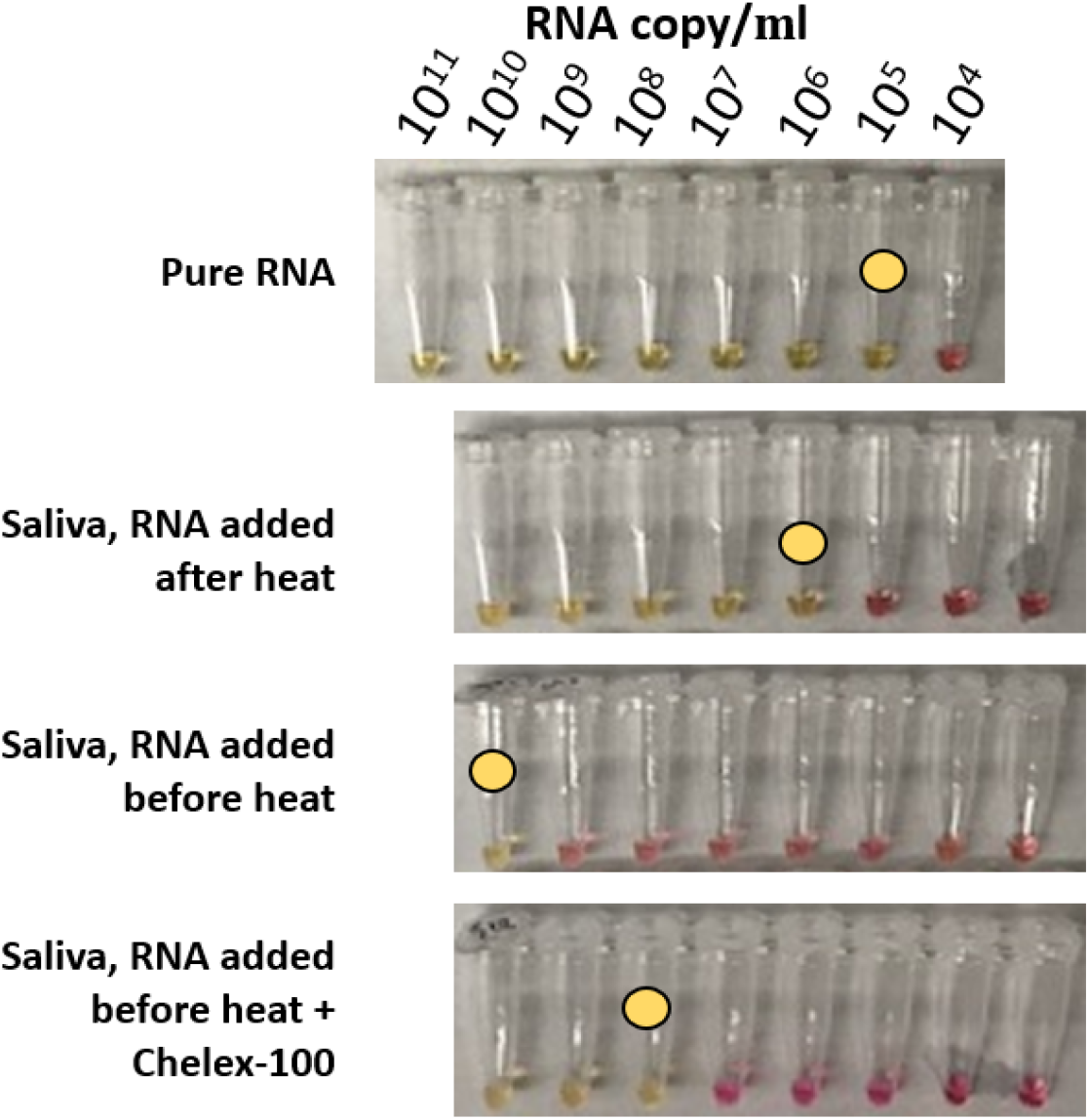
Chelex-100 partially protects RNA from heat induced degradation. In each tube, yellow colorimetric readout indicates RT-LAMP amplification. To prepare the samples, pure RNA was spiked into saliva at a 1:10 dilution, which is why the first panel is offset relative to the others. The limit of detection (LOD, yellow circle) of RT-LAMP on saliva samples is 10,000-fold lower if spiked RNA is added after heat inactivation (second row) than if it is added before (third row), indicating that a significant amount of RNA is being degraded as a consequence of the heat. Adding a chelating agent, Chelex-100, to the saliva before heat inactivation partially protects RNA from this degradation (fourth row).

### Optimization of Chelex-100 sample preparation

#### Temperature, incubation time, and Chelex-100 concentration affect sensitivity

Starting from a base protocol which used a 10 minute incubation at 95C with a 10% wt/vol Chelex-100 solution, we conducted one round of optimization by individually varying incubation time (Figure 2A), temperature (Figure 2B), and Chelex-100 concentration (Figure 2C), and compared the resulting limits of detection. The initial incubation time and temperature, 10 minutes and 95C, respectively, were near optimal. However, increasing Chelex-100 concentration from 10% to 30% wt/vol improved assay sensitivity 10-fold (Figure 2C). With the resulting optimal conditions, we achieved a sensitivity of 10^7^ copies per ml from human saliva (Figure 2C). However, SARS-CoV-2 genome concentrations in patient saliva range from 10^3^ to 10^9^ copies per ml^2,5^. Therefore, we sought additional optimizations before moving to clinical samples.

**Figure 2:**
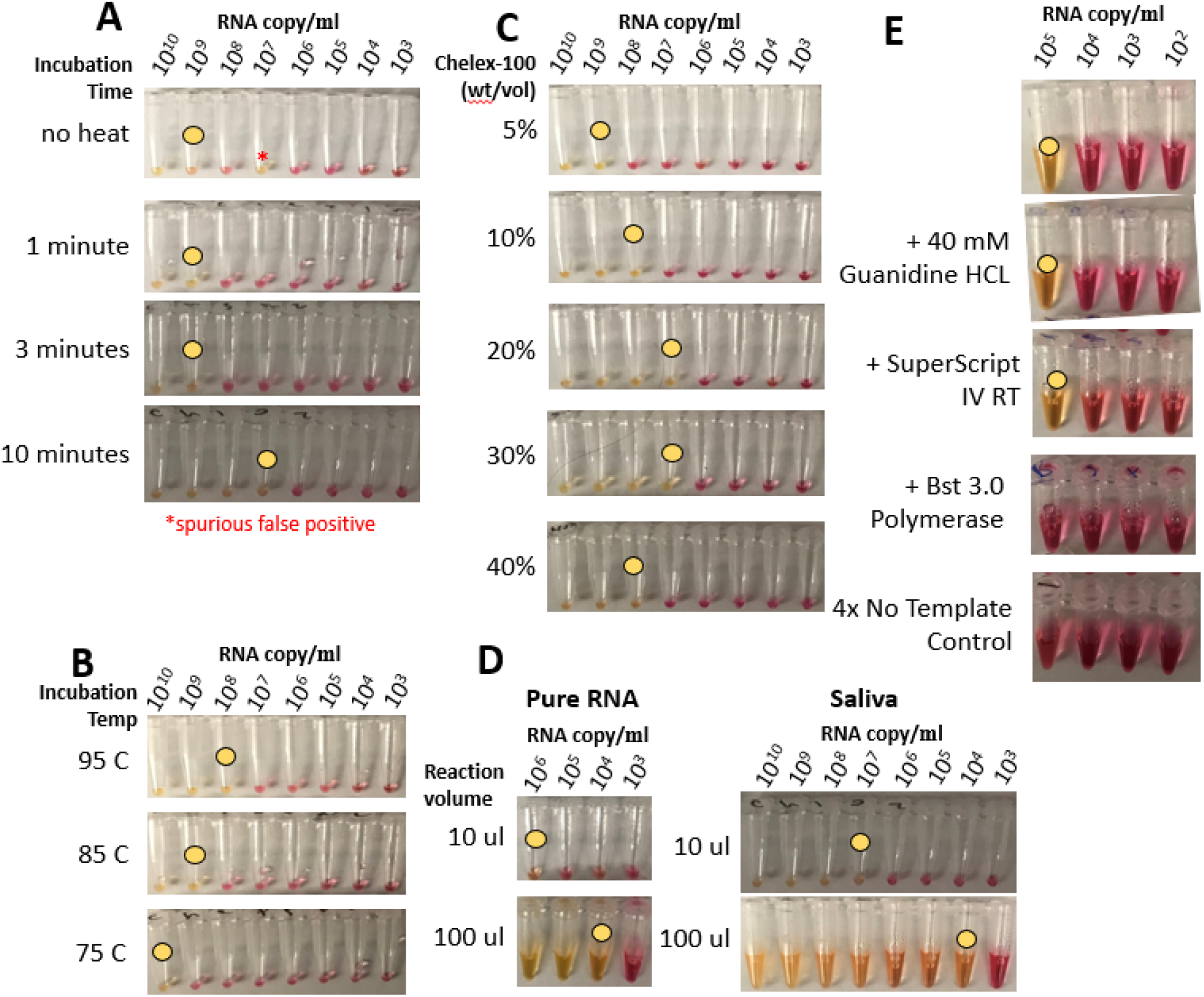
A Chelex-100 based sample preparation was optimized by varying parameters one at a time. Starting from an initial protocol where 10% wt/vol Chelex-100 was mixed 3:2 with sample and incubated for 10 minutes at 95 C, individual parameters were varied and the LOD (yellow circle) was compared. We found that (A) a 10 minute incubation time, (B) a 95C incubation temperature, and (C) 30% wt/vol Chelex-100 stock solution optimized LOD. Additionally, increasing the volume of the RT-LAMP reaction from 10 µl to 100 µl increased sensitivity by at least 100-fold (D), but increasing the volume further caused an increase in false positives (not shown). Based on reports in other publications, we also tried adding 40 mM guanidine HCL^22^, Superscript IV reverse transcriptase, and Bst 3.0 to increase sensitivity, however the LOD was unchanged by these additions (E). Based on these experiments, we called the final sensitivity at 10^5^ copies/ml.

#### RT-LAMP sensitivity scales with sample volume

Since RT-LAMP is an all-or-nothing reaction, its sensitivity should be proportional to the total number of template molecules. Intuitively, a larger volume at lower sample concentration should yield a similar result as a lower volume at higher sample concentration, assuming the same concentration × volume product. This would effectively increase sensitivity at higher volumes. To test this, we varied the RT-LAMP total reaction volume. Surprisingly, sensitivity increased super-linearly, rather than only linearly, as expected, over reaction volumes of 10 to 100µl. This occurred with both pure RNA samples and human saliva samples (Figure 2D). 100μl reactions with the Chelex-100 sample prep yielded LODs in saliva of 10^5^ copies per ml or better. Further increasing the reaction volume to 250 μl caused an increase in false positives (not shown), and was not pursued further.

#### Other optimizations failed to increase sensitivity beyond 10^5^ RNA copies per ml

40 mM Guanidine HCL was shown in other work to increase sensitivity of RT-LAMP^22^. Also, certain engineered polymerase and reverse transcriptase enzymes have been reported to be especially resistant to inhibitors, such as ThermoFisher Superscript IV reverse transcriptase and NEB BST 3.0 polymerase^31^. Starting from the optimized assay, we added each of these components to the reaction individually (Figure 2E). In each case, the resulting LOD was the same or worse than the base assay. Based on these results, and the sufficiency of 10^5^ copies/ml for recent studies of high frequency testing strategies, we decided to go forward with testing on clinical COVID-19 samples.

#### Chelex RT-LAMP measurement of unpurified clinical samples strongly correlated with qRT-PCR analysis of purified RNA

We evaluated the protocol on clinical throat and nose swabs, collected in universal or viral transport media (UTM/VTM) for analysis by the Virology laboratory at the Rambam Health Care Campus in Haifa, Israel. We compared qRT-PCR analysis of purified RNA to Chelex RT-LAMP analysis of the same samples without RNA purification. More specifically, we chose 31 positive and 31 negative samples that had been previously routinely analyzed in the laboratory by RT-qPCR (2019-nCoV detection kit, Seegene, CA, USA), with RNA extracted using either automated NucliSENSE easyMAG or using a magLEAD automated extraction platform. The RT-qPCR analysis produced C_t_ values ranging from 17 to 34. We then analyzed the original unpurified material for the same samples. For each sample, we mixed 60 µl of the sample with 90 µl 30% Chelex-100 solution, and heated to 95C for 10 minutes. Then, we added 10 µl of the resulting mixture to a 100 µl RT-LAMP reaction with the N2 primer set (Figure 3A). One negative control sample of sterile UTM was included in each set of samples (Figure 3A, label NC). Among the 31 negative samples, no strong positive outcomes were observed, although a few samples showed slight orange color (Figure 3A, arrowheads). By contrast, among the 31 positive samples, 27 showed strong color change. Among these, all 21 of the samples with C_t_<28 correctly showed positive results (Figure 3B). These results indicate a strong correlation between the Chelex RT-LAMP measurement on unpurified samples and the qRT-PCR analysis of purified RNA.

**Figure 3:**
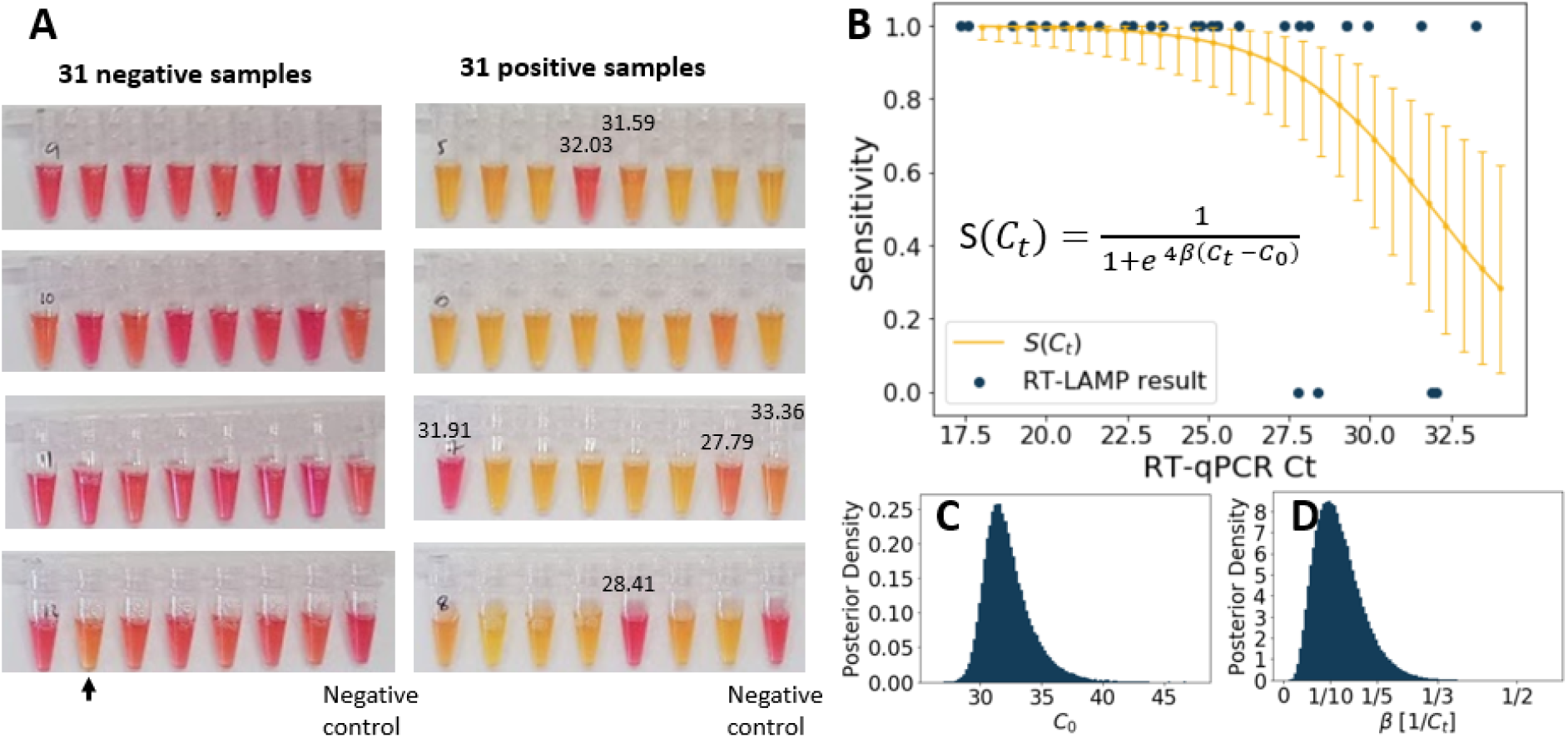
Clinical validation of RT-LAMP diagnostic on swabs in VTM. (A) 31 RT-qPCR positive and 31 RT-qPCR negative clinical nose and throat swab samples were collected and assayed for SARS-CoV-2 using the optimized protocol. 27/31 of the positive samples gave positive RT-LAMP results, with all 21 positive samples with C_t_ below 28 reading positive, and 0/31 negative samples giving positive results. (Note that some negative samples did show orange color, black arrow). (B) Sensitivity as a function of C_t_. The median estimated sensitivity and 95% confidence intervals at different C_t_. (C) Posterior distribution for C_0_. (D) Posterior distribution for β.

Defining sensitivity as the fraction of positive samples that were returned positive by the assay, we next asked whether we could identify a quantitative C_t_ threshold separating the regime with nearly perfect sensitivity from that where there is no detection. We assumed that the sensitivity of the diagnostic as a function of C_t_ could be described by a logistic model with two parameters: C_0_, defined as the C_t_ value where sensitivity S=½, and β, the slope of S at C_0_:

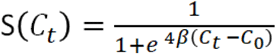

We fit the model using Bayesian inference (see Methods), yielding a posterior distribution with a median C_0_ estimate of 31.9 with a 95% confidence interval of [29.8, 36.0], and a median β estimate of 1/8.99 (95% confidence interval [1/19.9, 1/4.66]) (Figure 3B-D). More precise parameter estimates will require additional data. However, these inferences are consistent with the assay being highly robust for samples with C_t_ below 28 and sensitive enough to detect at least half of samples with C_t_ values of up to ∼32.

#### This protocol correctly identified 93% of positive saliva samples and 100% of negative saliva samples in a test of emergency room patients

As approved by the CMC Institutional Review Board, patients at Catholic Medical Center requesting COVID-19 tests volunteered saliva samples that were then marked with the positive or negative result of the PCR test. 20 negative samples and 14 positive samples were then assayed with the protocol developed in this paper (see Protocol), using the N1 primer set, which has similar sensitivity to the N2 primer set^32^. No negative samples yielded a positive readout after 45 minutes, however 13/14 positive samples did (93%, Figure 4). This confirms that this assay can be used as a simple, direct means to test saliva samples for SARS-CoV-2.

**Figure 4:**
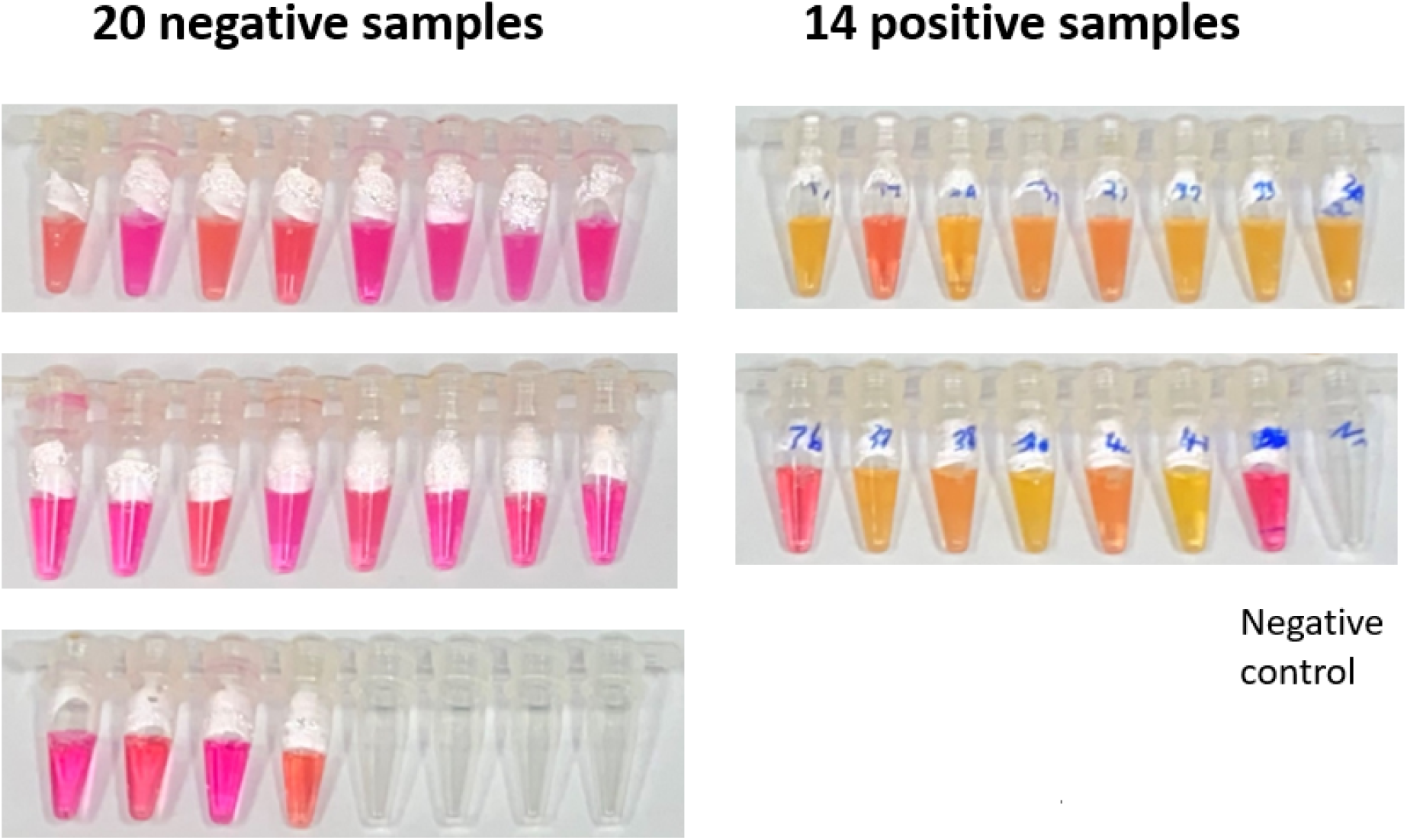
Clinical validation of RT-LAMP diagnostic on saliva samples. 20 negative saliva samples and 14 positive saliva samples were collected from patients at Catholic Medical Center in Manchester, NH and assayed with the Chelex-100 based RT-LAMP diagnostic. 0/20 negative samples gave a positive result, but 13/14 (93%) positive samples gave a positive result.

## Discussion

In this work, we optimized a SARS-CoV-2 detection protocol to prepare saliva and nasopharyngeal swab specimens for RT-LAMP without RNA purification. The use of high concentrations of Chelex-100 protected RNA from heat induced degradation, and a high temperature incubation inactivated inhibitors in the saliva. An increased volume of the RT-LAMP reaction allowed larger sample volumes in the reaction, which yielded increased sensitivity. Altogether, this protocol achieved a sensitivity of 10^5^ SARS-COV-2 RNAs / mL, robustly detected the virus in samples up to a Ct of 28, and was able to detect the virus in samples with Ct up to 32, without false positives in negative samples.

This protocol enables the implementation of high throughput population screening. Saliva collection is safer, easier, and less invasive than swab collection. Since Chelex-100 is a non-hazardous reagent, it can be included in self-collection kits. Once samples have been collected, they can be quickly heat inactivated, and processed in parallel on PCR plates, allowing many reactions to be conducted at a time. By contrast, many alternative diagnostic protocols require uncapping of samples, mixing of additional reagents into samples, and other kinds of sample handling such as centrifugation to purify RNA or otherwise prepare the sample for the assay. By making this sample handling unnecessary, this sample prep method is safer, faster, and higher throughput.

There are still issues to be worked out. A limitation of this protocol is that the increased volume of the assay leads to a reaction cost of ∼$7. This cost can be potentially reduced by reducing the volume to the 25 µl to achieve ∼$2 per reaction, albeit with reduced sensitivity. Additionally, use of open source enzymes^26,33^ and pooling the samples could further reduce costs. As the volume of LAMP reaction increases, false positive results seem to increase in frequency which manifested as a few negative samples turning orange in our experiments presented here. These problems could potentially be mitigated by the inclusion of UDG/UTP in the LAMP reaction, which others have used successfully to reduce false positives^26^. These problems could also be reduced by running reactions with 2 different primer sets, which should increase certainty of a positive or negative result.

With the current surge in Covid-19 across the US and the world this technology could prove very helpful to fill the void for rapid, affordable, easily deployable, widely distributed testing.

## Data Availability

All data from this study can be obtained by emailing the first author.

## Acknowledgements

This project was supported by the Caltech Merkin Institute for Translational Research and the Israel Science Foundation (grant No. 3633/19) within the KillCorona – Curbing Coronavirus Research Program. James Flynn provided his own funds to conduct the study at Catholic Medical Center.

## Methods

### Bayesian Inference

The Metropolis-Hastings algorithm was used to sample parameters from the posterior distribution of β and C_0_, using the logistic model

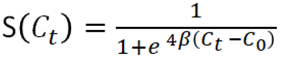

Priors were constructed by analyzing the expected scale of the parameters. Since it was unlikely that this LAMP assay did significantly worse or better than PCR, a Gaussian prior was chosen for C_0_, with mean 30 and standard deviation 10. For the β prior, we noted that the derivative of the sensitivity function at C_t_ = C_0_ is -β and that we expected the sensitivity to drop from around 1 at C_t_=28 to around 0 at C_t_ = 35, which would imply the scale of the slope is on the order of magnitude of 1/7. Thus, the β prior was chosen to be gaussian with mean 1/7, with standard deviation equal to 2/7, to allow for some flexibility. Both priors were restricted to the positive real line. Transition probabilities for C_0_ and β were gaussian with mean zero and standard deviation of 1 and .1, respectively. 1 million points were sampled from the posterior distribution, and the .05, .5, .95 quantiles for both β and C_0_ were computed.

### Protocol

#### Materials

- Chelex-100 (Bio-Rad #1421253)
- WarmStart® Colorimetric LAMP 2X Master Mix (NEB #M1800S/#M1800L)
- Nuclease free water (VWR #10220-398)
- N2 primers:
- N1 primers:

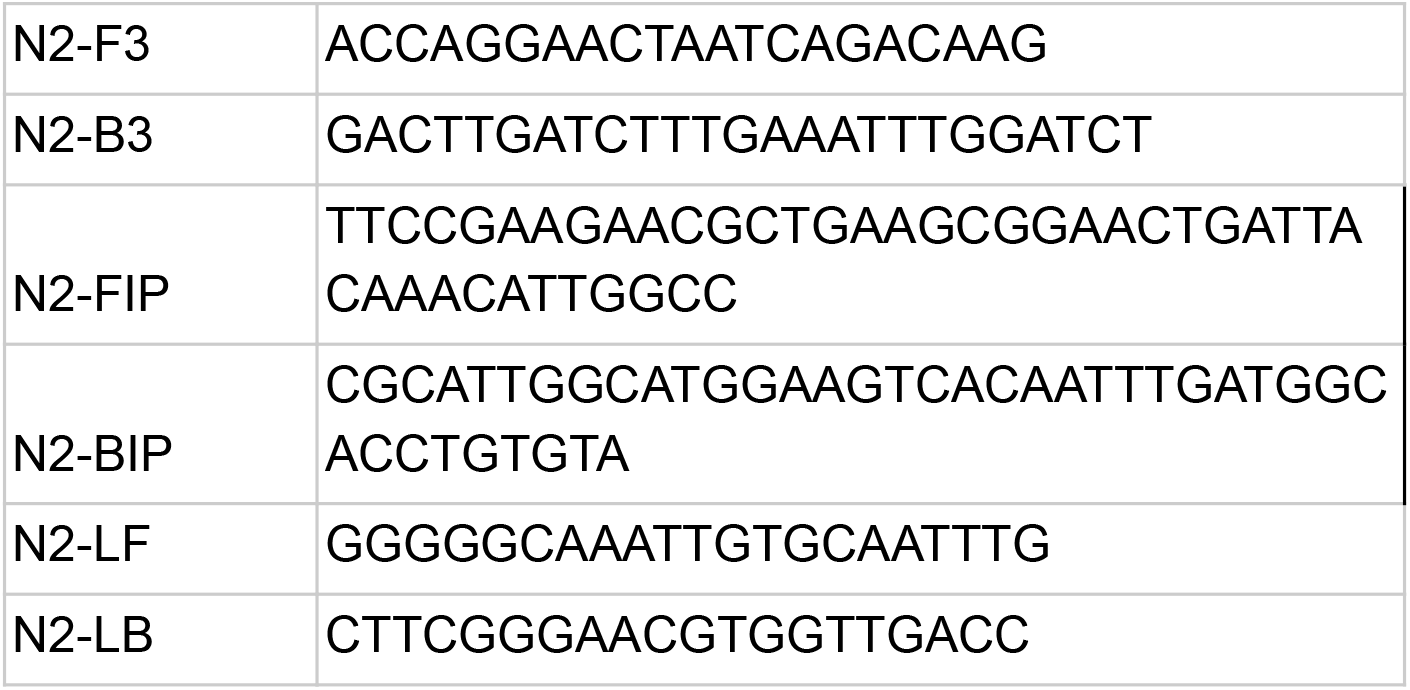

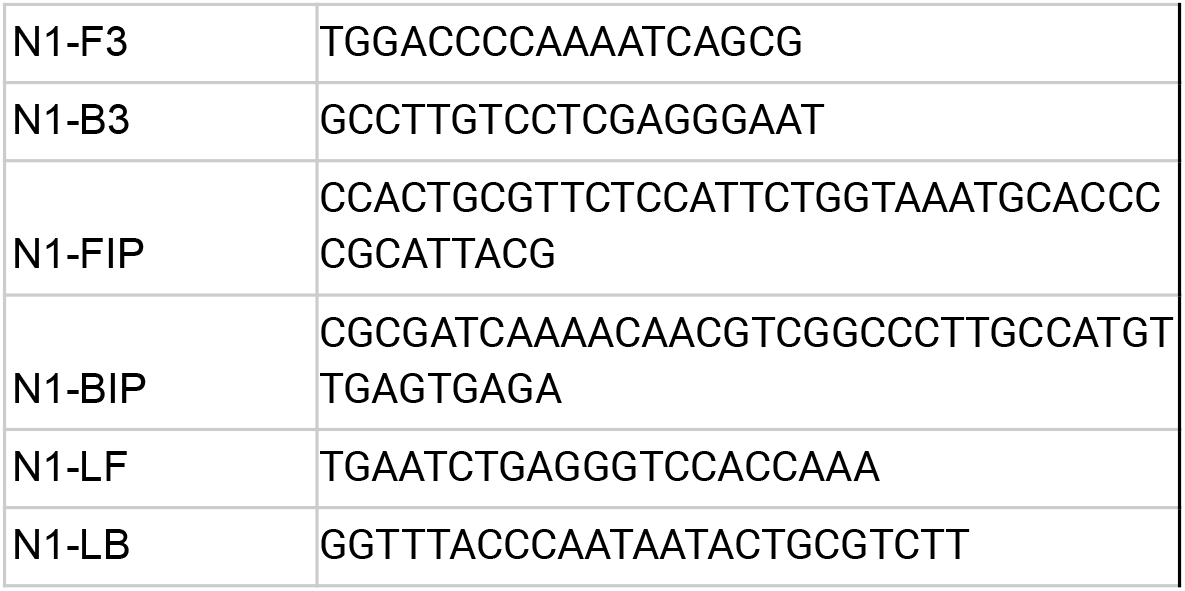

#### Important Note

It is recommended that these primers (both N1 and N2) be PAGE purified for the highest sensitivity and lowest off-target samplification.

### Prepare 30% wt/vol Chelex stock solution

1. Add 4.5 g Chelex-100 to a 15 mL tube
2. Fill with deionized water up to 15 mL
3. Shake to mix until no white clumps are visible. The final solution should look like a milky slurry that settles to the bottom of the tube relatively quickly (∼ 1-3 mins).

### Prepare Primer Master Mix

In a PCR tube, mix:

1. 56 µl nuclease free water
2. 16 µl N2-FIP
3. 16 µl N2-BIP
4. 4 µl N2-LF
5. 4 µl N2-LB
6. 2 µl N2-F3
7. 2 µl N2-B3

This gives 10 reactions for a 100 µl volume LAMP reactions. For more reactions, these numbers can be scaled proportionally.

### Sample preparation

For 100 µl of fresh saliva/swab in VTM

1. Shake Chelex-100 stock solution vigorously to resuspend the resin (because Chelex is quick to settle, it should be reshaken every 3 samples or so if they are being done in succession)
2. Add 150 µl of the resuspended 30% Chelex-100 stock solution to the 100 µl saliva, pipette up and down to mix thoroughly.
3. Incubate samples in a 95 C wet bath (or thermocycler or aluminum 96-well plate) for 10 minutes.
4. While the Chelex-saliva solution is heating, prepare a mixture of
  - 50 µl Warmstart Colorimetric LAMP Master Mix (thawed on ice)
  - 10 µl N2 primer master mix
  - 30 µl nuclease free water on ice for each sample.
5. After 10 minutes, remove Chelex-saliva solution from heat, place on ice for 2 mins.
6. Spin Chelex-saliva solution in a PCR tube mini centrifuge for a 5 second pulse.
7. Pipette 10 µl from the **top** of the Chelex-saliva supernatant into the Lamp mixture, final volume should be 100 ul.
8. Heat the final lamp mixture at 65 C for 30 minutes.
9. Readout: yellow = positive, pink = negative.

